# Comparison of contact diaries and wearable proximity sensors in measuring household contacts in low- and middle-income countries

**DOI:** 10.1101/2025.10.07.25336904

**Authors:** Machi Shiiba, Moses C Kiti, Obianuju Aguolu, Noureen Ahmed, Charfudin Sacoor, Ivalda Macicame, Edgar Jamisse, Corssino Tchavana, Orvalho Augusto, Americo Jose, Migdalia Wamba, Nilzio Cavele, Azucena Bardaji, Herberth Maldonado, Rajan Srinivasan, Venkata Raghava Mohan, Momin Kazi, Raheel Allana, Fauzia Malik, Benjamin A Lopman, Saad B Omer, Kristin N Nelson

**Author notes:** Corresponding author: Machi Shiiba.

## Abstract

**Background:** Respiratory and enteric infections remain a high burden worldwide. The spread of these infections is driven by person-to-person interactions, or ‘contacts’. Contact diaries have been widely used to collect information such as the number of contacts a person has throughout the day, and characteristics of each interaction that may be relevant for disease transmission. Wearable proximity sensors have been used to complement data from contact diaries, and in particular to capture contacts among children who are not able to report their contacts. Studies that have compared contact diary and wearable proximity sensors are mostly in high-income countries or conducted in small populations in low- and middle-income countries (LMICs). We aimed to compare how effectively the two methods capture contacts relevant for disease transmission in LMICs’ household settings.

**Methods:** We conducted a population-based study and collected social contact data with contact diary and wearable proximity sensor in four LMICs – Guatemala, India, Mozambique, and Pakistan. We compared the number, concordance, and duration of contacts captured by the sensors and the reported in the contact diaries.

**Results:** 421 participants documented a total of 1189 unique contacts on contact diaries, and recorded 1248 unique sensor contacts. The mean number of unique contacts was higher in the contact diary than recorded by the sensors in India, lower in the contact diary than recorded by the sensors in Mozambique and Pakistan, and similar in Guatemala. In all countries, more than half of contact pairs were reported by both contact diary and sensors, though this varied by site with 56% of contact pairs reported by both methods in Mozambique and 87% of contact pairs in India. The duration of contacts reported by concordant contact pairs was longer in contact diary compared to the duration captured by sensors in all four countries.

**Conclusions:** This study demonstrated that both contact diaries and wearable proximity sensors have unique features in measuring social contacts in LMICs, suggesting that they are complementary. Future work should consider the characteristics of each method in understanding person-to-person interactions relevant for infectious disease transmission and implement suitable options for the scope of their study.

## Background

Lower respiratory and enteric infections are the leading causes of mortality due to infectious diseases, especially among children under 5 years of age worldwide (1, 2). The spread of infection is driven by person-to-person interactions, henceforth referred to as ‘contacts’. Data on contact patterns are an essential component in mathematical models for understanding age-specific infection dynamics and estimating the effectiveness of intervention strategies (3, 4).

The standard method used to collect data on contacts is a survey, or ‘contact-diary’, henceforth called ‘diary’. Participants complete information on each interaction over a short time period (e.g. one day), including the age, sex, duration, and location of the interactions. While diaries can collect rich information on contacts, they are subject to recall bias, such as omitting non-salient contacts and inaccurately estimating contact duration. For example, people may forget to report routine interactions with household members such as spending time with close relatives in commonly visited areas.

Wearable proximity sensors, henceforth called ‘sensors’, have been introduced to complement contact-diary reports. Studies using sensors to capture face-to-face contacts have been conducted in hospitals, schools and conferences; mostly in high-income countries (5-9) and household settings in low-and-middle-income-countries (10, 11). One advantage of the sensor is that it does not require participants to remember each contact (12). Additionally, sensors can be deployed autonomously in large group settings, including people who cannot self-report contacts, especially young children and illiterate adults. They also provide highly temporal granular data and over longer duration compared to diaries (5, 10, 13, 14). However, this method has several limitations, key being that they can only capture contacts of those who are wearing similar sensors as they rely on the signal exchange. This limits the usage of sensors to closed populations who are required to wear it throughout their daily activities (12, 15). Additionally, sensors do not distinguish between contacts that may be more or less likely to lead to transmission, since they do not directly capture whether a contact was physical, or the outdoor environment in which it occurred (16). Lastly, maintaining data quality can be challenging as it is dependent on participants’ compliance and the proper functioning of all devices.

There are several studies that have compared the usability and reporting characteristics of diaries and sensors. These studies were conducted in high-income settings targeting school and conferences in the US, France, and Germany (14, 15, 17-19). Participants in 4 out of the 5 studies underreported short duration (lasting < 5 minutes) contacts (14, 17-19). This suggested that participants often forgot to report short interactions in diaries. These discrepancies arose partly from differing definitions of contact used by diaries and sensors. In these studies, sensors were designed to capture contacts when two or more individuals wearing sensors hanging on their chest were face-to-face, within 1.5 meters, and with no obstructing objects with 20 seconds temporal resolution. Contacts in a diary were defined as close face-to-face interactions in one study, and physical touch or exchange of 10 or more words in another study (14). However, even when definitions of contact were standardized across the methods, sensors were more sensitive to short-duration contacts. Importantly, these are more likely to be the contacts where people were co-located in close proximity but did not engage in any conversation – contacts which may still be highly relevant for disease transmission (17). When comparing the duration of contacts recorded in diaries to those measured by sensors, participants tended to report longer duration contacts in the diaries. This indicates a tendency to overestimate how long interactions lasted when reporting in diaries (14, 15, 17).

One study reported that although the characteristics of contacts such as the average number of nodes and links were different in diaries and sensors, the overall structure of contact network such as the strongest links and clusters, in other words, more likely to transmit infections, are captured by both methods (17). Additionally, another study found that although the two methods reported different individual contacts, when the contact matrices from diary and sensors are used in a model, they resulted in a similar attack rate for influenza like illness in school settings (18). All these studies were conducted in high-income countries and mostly in school settings. However, households are important in the transmission of respiratory and enteric infections, yet poorly studied (20). Previous studies in low- and middle-income countries using sensors involved small household samples and the results were difficult to generalize to larger populations (13, 21). Therefore, it is crucial to study how sensors perform in household settings in LMICs in a larger population to inform the collection of accurate contact data in low- and middle-income countries.

Our study aimed to validate how well the data from diaries and sensors match to each other and thus, examine to what extent data from the two methods reflect social behavior relevant for disease transmission in low- and middle-income countries’ household settings.

## Methods

### Data collection procedures

The GlobalMix study aimed to characterize social contact patterns in low- and middle-income countries. One of the main objectives was to characterize the social contacts of infants with their household members using sensors. A detailed study design is described in a previously published paper (22). Briefly, the study was conducted in one rural and one urban site each in Mozambique, Guatemala, India and Pakistan. In each site, we recruited 63 households with a child less than 5 years old (index child) to wear sensors. The selection of index child was proportional to three age groups (<6 months, 6-11 months, 1-4 years old) with oversampling of infants less than 6 months old as their data were not usually available from prior studies and their contact behavior was of particular highest interest. At least 75% of household members in India, Mozambique, and Pakistan were required to participate and no such threshold was set for Guatemala (Data collection procedures were tailored to each site). We used diaries and sensors to measure human interactions relevant to infectious disease transmission, especially those of young infants (22).

Contact diary: Participants were trained to fill in a paper-based contact diary for two days to record who they interacted with and what were the characteristics of those contacts such as contacts’ age, gender, relationship to the participant, contact duration and location. For participants who were unable to fill in the diary themselves (e.g. infants and illiterate adults), a shadow filled in the diary on their behalf. We defined a diary contact as a two-way conversation with three or more words exchanged in the physical presence of another person or directly touching someone or clothes they were wearing (e.g. handshake, hug) (22).

Wearable proximity sensor: Members of participating households were instructed to wear a sensor for seven days that included the two days they completed the diary. Participants were instructed to wear the sensors on their chest at all times when in the house, from the time they woke up until the time they went to bed. We asked participants to position the sensor around their chest by either wearing it on a neck lanyard, placing it in the front pocket of their shirt or blouse, or attaching it to their clothing with a clip. The sensors exchanged data packets when two or more participants wearing a sensor on the chest area were face-to-face within 1-1.5 meters of each other. Data packets contained, among others, the sensor identification number (ID) and transmitted and received RSSI (an indicator of signal strength) to estimate the proximity between sensors. We defined a sensor contact via sensor data as occurring between two or more individuals if their sensors exchanged at least one data packet over a 20-second interval (22).

At the Pakistan rural site, we recognized multiple households in a community interacted and spent time together throughout the day, therefore, we treated them as one community and assigned the same household ID to capture their daily interactions. For this analysis, we required each household to have unique household ID to identify unique contacts in diary, therefore, we excluded Pakistan rural site from this analysis. All the results in this paper for Pakistan include only urban site.

### Descriptive Statistical analysis

We identified contacts reported in diary by matching contact age group, contact sex, and contact’s household ID to participant age group, participant sex, and participant household ID as most of the household contacts were theoretically also participants of this study. If we could not identify a definite match between a contact in the diary and a participant, or had multiple potential matches, we did not include those contact data for this analysis.

We also removed participants and their contacts from the sensor data with missing sensor IDs, incorrect or inconsistent household or personal identifiers, or if the data suggested that participants had not worn the sensor. In the final dataset, we included households with at least two household members and households that had sensor data available for all members, which we refer as analytic participants in this paper.

We compared the demographic characteristics of participants who participated to wear sensors in the overall GlobalMix study with the participants who were included in our analysis. To assess the agreement between the two methods, we compared the mean number of unique contacts between diary and sensor. Paired t-test was used for testing the statistical significance of the difference in means of contacts from the two methods in each country. To assess the differences in contacts captured by each method, we reported the number of contact pairs (unique pairs of participant and contact) recorded in the diary only, detected by sensor only, or captured by both methods. Then, we reported the proportion of the number of contact pairs recorded in diary only, detected by sensor only, or captured by both methods by participant age, participant sex, and household member relationship to examine whether there were socio-demographic factors that made contacts reported more in one of the methods. We also conducted logistic regression analysis to examine the association between these demographic characteristics and contacts reported by each method. Two models were fitted per country: one with the outcome defined as contacts reported only by diary (vs. sensor or both), and another with the outcome defined as contacts reported only by sensor (vs. diary or both). Predictor variables were the same variables used for the descriptive analysis above.

Lastly, we compared the duration of contact reported for contacts captured by both methods. Although participants recorded both household and non-household contacts in the diary, we restricted this analysis to household contacts, which were measured by both methods.

### Data availability

The data, analysis code, and survey questionnaires for this analysis is available at https://github.com/lopmanlab/GlobalMix-sensor-analysis.git

### Ethical Approvals

Informed consent was obtained from all participants 18 years old or older for themselves to participate in the study. For all individuals younger than 18 years, informed consent was obtained from their parents and guardians for their children to participate in the study. Assent was also obtained from individuals 12-17 years old for themselves to participate in the study. Additionally, consent for household heads were obtained to give consent for themselves and all members of their household to participate in the study. This study was conducted in accordance with the Declaration of Helsinki, and was approved by Emory University Institutional Review Board (approval number 00105630), Yale University (reliance agreement approval number 2000026911), UT Southwestern (reliance agreement approval number STU-2023-1107), and review boards at Christian Medical College Vellore (Approval #12065), Universidad del Valle de Guatemala (Protocol #223-12-2020) and the National Ethics Committee from the Ministry of Public Health and Social Assistance in Guatemala (Protocol #31-2020), Manhiça Health Research Institute Internal Scientific Committee (CCI/03/2020) and Internal Ethical Review Board (initial approval CIBS-CISM/011/2020), and The Aga Khan University (Approval #2022-7912-23648).

## Results

There were 1880 participants in the GlobalMix study (n = 342 in Guatemala, n = 737 in India, n = 579 in Mozambique, n = 222 in Pakistan). We excluded participants with no valid sensor data. This included (1) missing or unidentifiable data of the participants due to loss of sensors or sensor IDs or incorrect household and personal identifiers; (2) sensors recorded no data or inconsistent data such as 48 hours continuous recording suggesting the participants not wearing sensors but put them all together at the same place; (3) person living alone so no contacts should be counted as a household contact. After filtering out these participants, 439 participants (n = 61 in Guatemala, n = 211 in India, n = 94 in Mozambique, n = 73 in Pakistan) remained. Then, 421 participants (n = 53 in Guatemala, n = 211 in India, n = 88 in Mozambique, n = 69 in Pakistan) remained after filtering out the participants whose household contacts in diary were not identifiable. Those 421 participants reported total of 1189 unique diary contacts (n = 101 in Guatemala, n = 694 in India, and n = 226 in Mozambique, n = 168 in Pakistan) and 1248 unique sensor contacts (n = 100 in Guatemala, n = 636 in India, n = 280 in Mozambique, n = 232 in Pakistan) over two days. (Supplemental figure 1)

Compared to all recruited participants, the distribution of participants’ age and sex were similar to those with available sensor and diary data. We presented results of the latter which made up our analytic dataset. The participants included in this analysis had lower proportion of rural population in Guatemala and Mozambique (0.28 vs 0.54 in Guatemala, 0.39 vs 0.54 in Mozambique) while it was the opposite in India (0.65 vs 0.51), and this was not applicable to Pakistan as only urban site was included. (Table 1, Figure 1)

**Table 1:**
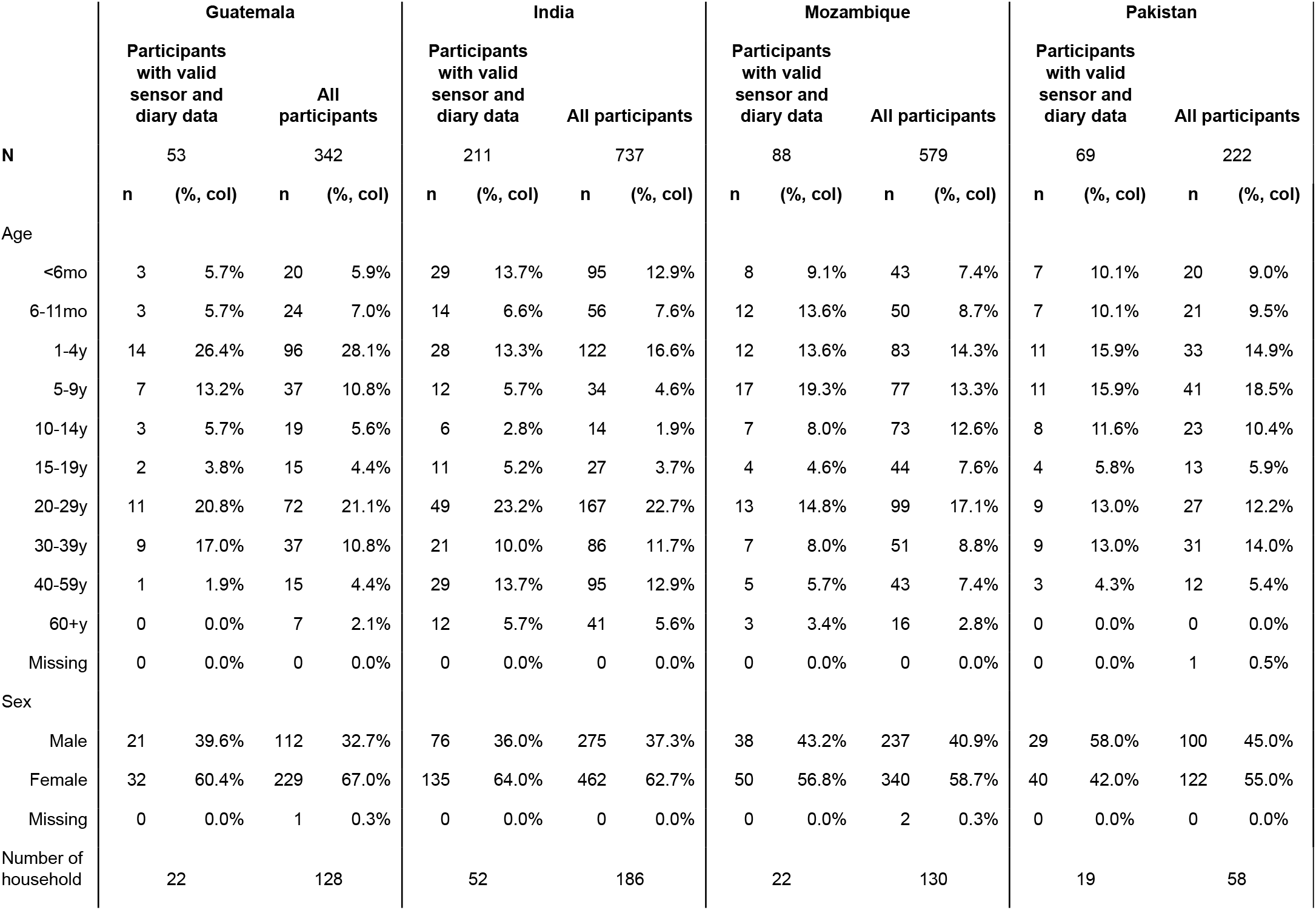
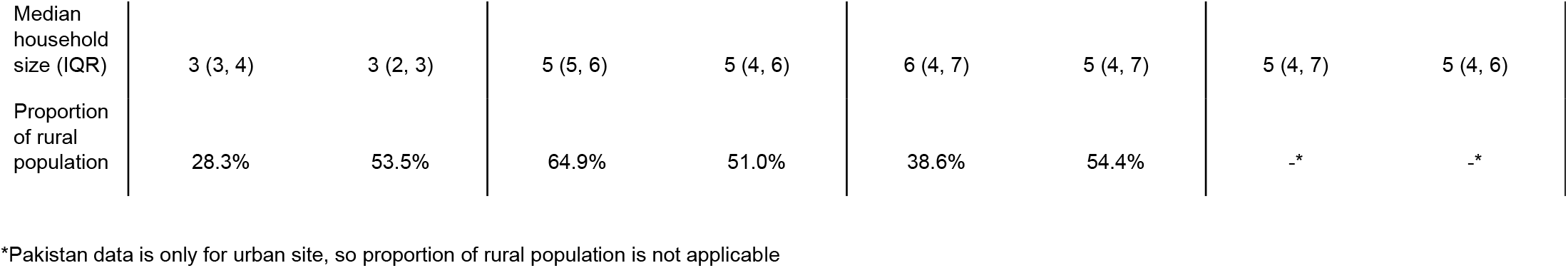
Characteristics of study participants and household by country.

**Figure 1.**
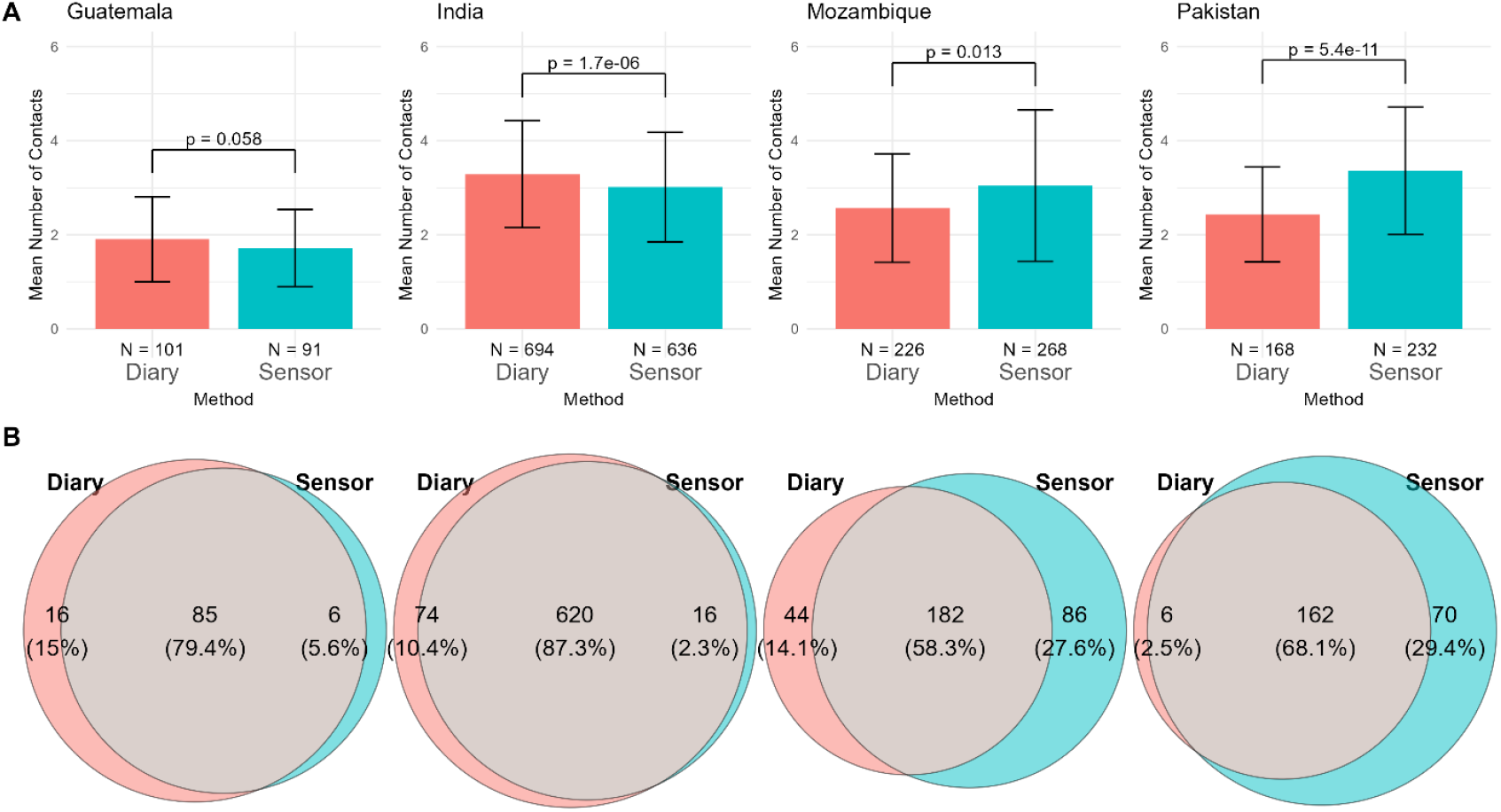
Comparison of number of unique contacts in contact diary and wearable sensor methods. Panel A shows the mean number of unique contacts over two days in contact diary and wearable sensor in each country. Bars show 95% confidence intervals. The values at the bottom of the bar shows the total number of unique contacts recorded in each method. The values on top of the bars show a p-value from the paired t-test on the mean number of unique contacts. Panel B shows the number and percentages of pairs of contacts recorded in either contact diary (red), wearable sensor (blue), or in both methods (grey).

We found variation by country in the overlap between contacts reported by each method. India had the highest proportion of contacts reported via both diary and sensor (87%), followed by Guatemala (79%), Pakistan (68%), and Mozambique (56%). The proportion of contacts that were recorded by sensor only was substantially higher in Mozambique and Pakistan compared to the other two countries (28% in Mozambique, 29% in Pakistan, 6% in Guatemala, and 2% in India). The mean number of unique contacts reported in diary across sites were 2.8 (SD: 1.2) and the number captured in sensor was 2.9 (SD: 1.4). The mean number of unique contacts reported in the diary was higher compared to the number captured using sensors in India (mean: 3.3 contacts, 95% CI: [3.1, 3.4] by diary, mean: 3.0 contacts, 95% CI: [2.9, 3.2] by sensor, p < 0.05) while the number of unique contacts in diary was lower than sensors in Mozambique (mean: 2.6 contacts, 95% CI: [2.3, 2.8] by diary, mean: 3.0 contacts, 95% CI: [2.7, 3.4] in sensor, p < 0.05) and Pakistan (mean: 2.4 contacts, 95% CI: [2.2, 2.7] by diary, mean: 3.4 contacts, 95% CI: [3.0, 3.7] by sensor, p < 0.05). Mean contacts were similar between methods in Guatemala (mean: 1.9 contacts, 95% CI: [1.7, 2.1] by diary, mean: 1.7 contacts, 95% CI: [1.5, 1.9] by sensor, p = 0.06). (Figure 1)

The pattern in the proportion of contact pairs reported by both diary and sensor, by participant age, varied by country. In Guatemala, the youngest (<6mo) and oldest age (40-59y) groups had relatively high proportion of contacts that were only captured by sensors compared to other age groups. In India, most contacts were reported by both methods and some reported by diary only, and this did not differ by age groups. In Mozambique, proportion of contacts reported by diary only was higher in 6-11 months and 60+ years old group than those by sensor only while most other age groups were the opposite with more than 25% of contacts reported by sensor only. In Pakistan, certain age groups (5-9y, 15-19y, and 40-59y) had higher proportion of contacts detected by sensor only compared to other age groups although the patterns remained relatively similar across all age groups. The patterns did not differ significantly by participant sex in all four countries. The proportion of contact pairs reported by each method by contact’s relationship to participants did not show clear patterns and could not make a definitive conclusion. (Figure 2, Supplemental Figure 2)

**Figure 2.**
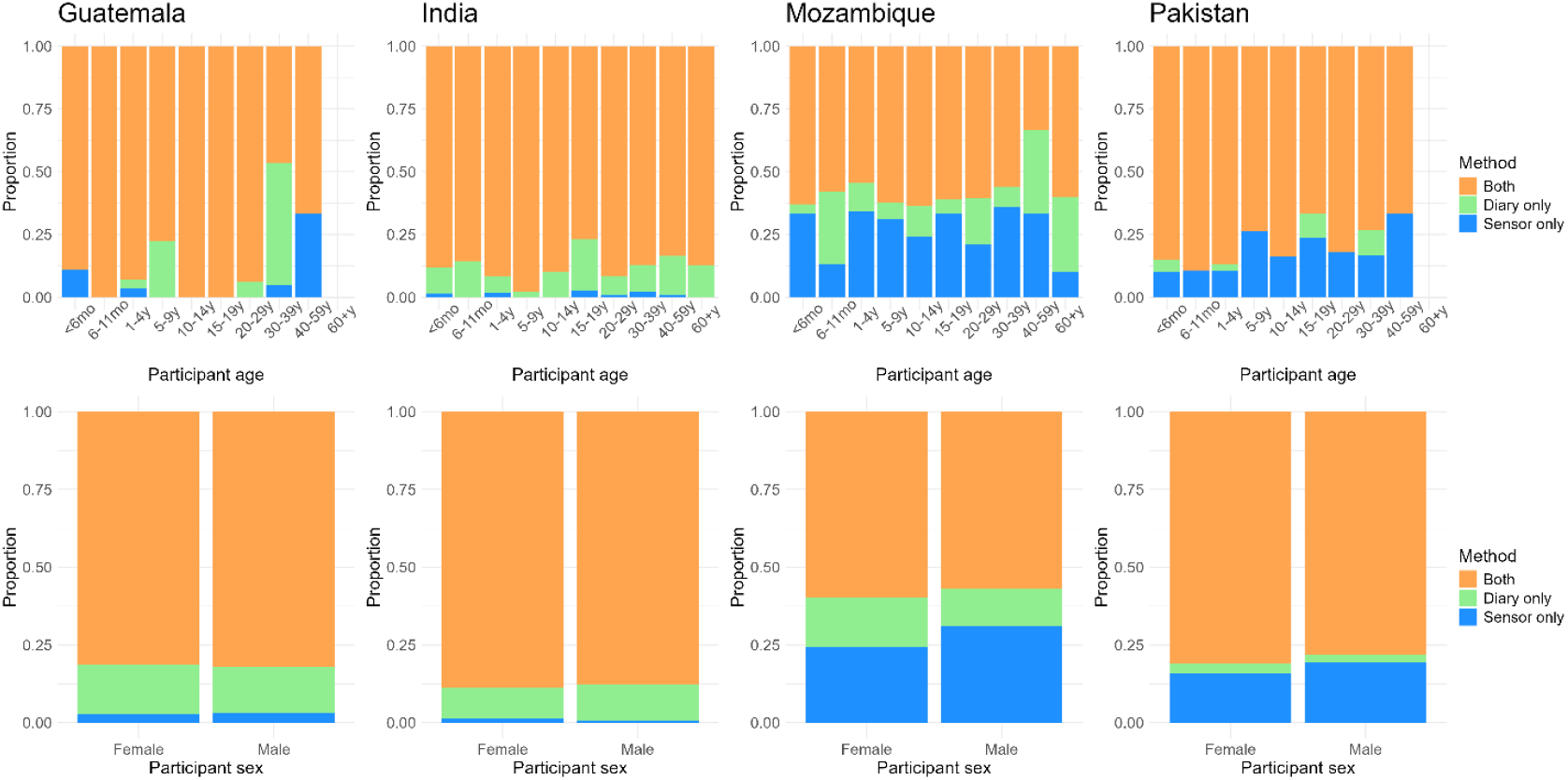
Comparison of contacts measured in two methods by participant demographics. The first row shows the proportion of the number of contact pairs reported by both methods, diary only, and sensor only by participant age and country. The second row shows the same proportion by participant sex.

Regression analysis results indicated additional older contacts were likely to be reported by diary (Odds Ratio > 1) in some countries, which were not the case for sensors. Overall, there were no clear patterns of contacts characteristics that were more likely reported by diary or captured by sensor across four countries. (Supplemental Table 1)

In all four countries, participants reported in the diary longer duration of contact than was captured in the sensor data. The proportion of contacts lasting longer than one hour were higher in diary compared to sensor (99% in diary and 46% in sensor in Guatemala, 81% in diary and 40% in sensor in India, 100% in diary and 53% in sensor in Mozambique, 100% in diary and 32% in sensor in Pakistan). In India and Pakistan, there were substantially higher proportion of shorter contacts (less than 5 minutes) detected by sensor compared to the other two countries (33% in India, 36% in Pakistan, 16% in Mozambique, 7% in Guatemala). (Figure 3)

**Figure 3.**
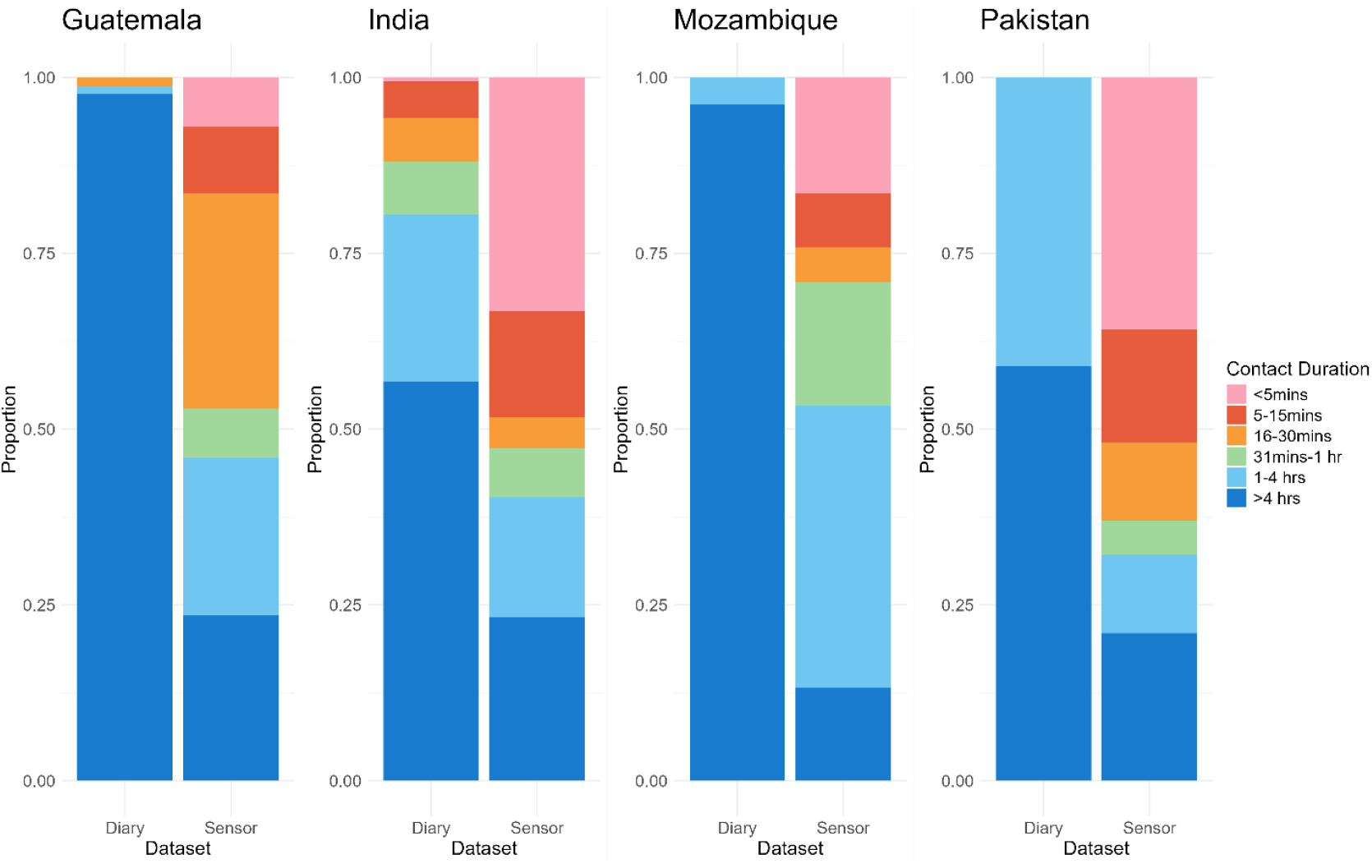
Proportion of contact duration by measurement method. Proportion of contact duration reported by diary is shown on the left bar and captured by sensor is shown on the right bar for each country. Contact duration categories were taken from the categories used in the contact diary. Sensor’s contact duration was calculated from seconds to match the categories.

## Discussion

We compared the characteristics of household social contacts reported in a contact diary and captured by wearable proximity sensors across seven sites in four low and middle-income countries. The methods did not perform uniformly, and there was not one that consistently collected a higher number of contacts: participants in India reported a higher number of contacts in the diary than sensors, but diary reported lower number of contacts in Mozambique and Pakistan, and the two methods recorded similar number of contacts in Guatemala. The patterns of contacts reported by method, stratified by participant age group, differed by country; no clear pattern was observed when stratified by participant sex. However, a consistent finding was that the duration of contacts reported in the diary was longer than ones captured by sensors for all four countries.

Inconsistency in which method measured a higher number of contacts across countries indicate that the validity of these two methods may differ by context, and the data they capture may be complementary rather than overlapping. Previous studies comparing diary and sensor methods of collecting contact data have reported mixed results depending on the setting. Two studies in a US school setting (including from kindergarten to high school) reported higher numbers of contacts measured by sensors compared to a diary. This was attributed to sensors capturing contacts of short duration (<5 minutes), while diary missed those shorter contacts. (18, 19). Given the focus of our study on household contacts, contacts tended to be long in duration, which could be a reason for inconsistency in the patterns of mean number of contacts with previous studies. Additionally, recall bias may be less likely in our study as we focused on household contacts that are likely repeated and longer compared to other studies focusing on non-household settings with fleeting non-familiar contacts. Other studies found that the mean duration of contacts was longer in diary than sensors because participants tended to forget, and fail to report, shorter duration contacts in diary (14, 17). Although our analysis of duration of contacts compared the same contact pairs between two methods, which is slightly different from the previous studies, we could say that the trend of participants reporting longer contacts in diary than sensors followed the results of previous studies. This suggests that people overestimated the time they spent with someone. It is also important to note that the differences we observed in contact duration between the diary and sensor methods in our analysis could be due to differences in how participants recognized contacts in diary and how sensors captured contacts. Sensors captured contacts only when participants were face-to-face within 1-1.5 meters during the day. However, in the diary, participants may have reported contacts when they talked to other people in the same room but not necessarily facing each other, or some participants may have reported sleeping with other household members as contacts.

Our analysis showed that participant characteristics were similar between all participants and the analytic participants. Therefore, we postulate that the results are representative of the overall GlobalMix study participants and their contact characteristics. However, here are some limitations to note. One, there were many missing and inconsistent sensor data, due to the technical challenges of using sensors in the household which led to the removal of more than 75% participants from which sensor data was initially collected from the analytic dataset. Poor quality sensor data could have been attributed to multiple factors, including participants forgetting to wear sensors, being unwilling to carry and wear sensors, losing sensors, and improper wearing of sensors. These data were omitted for the analysis. However, it is important to note that the contact characteristics (contact age, contact sex, contact duration) of the overall and analytic data did not differ greatly. Although our study performed extensive qualitative work to understand community perceptions of sensors and to tailor the study protocols which involved sensors to each context, compliance remained a challenge. Additionally, some of the data collection especially in Mozambique and Guatemala was during the COVID-19 pandemic which may have affected participants’ daily activities and compliance in this study. Another limitation of this study is that some of the contacts reported in diary were not identifiable; the largest proportion of unidentifiable diary contacts was in Mozambique. This most likely caused the number of contacts reported and captured by both methods to be lower than the other countries. Additionally, it resulted in filtering about half of the contacts in Mozambique when comparing the duration of contacts. Lastly, while the sensors have been used in measuring contacts in multiple studies, they may no longer be the most advanced or accurate technology to measure contacts. Other technologies such as smartphone applications, GPS tracking on phones or smartwatches could potentially provide higher compliance among participants although the practicality, especially in terms of cost and storage of larger data, needs to be considered (23, 24). Additionally, these devices capture the information that some people are co-located rather than their interactions. For example, two people could be in the same room but not facing each other or could be in the same building with a wall between them which are not relevant for infectious disease transmission. Therefore, the sensor was a suitable choice for this study population for the scope of our study.

## Conclusions

This study highlighted the relative performance and characteristics of diary and sensor in measuring human social interactions in diverse settings in LMICs. It demonstrated that both contact diaries and wearable proximity sensors have unique strengths, suggesting they are complementary measurements for capturing social contact patterns. The result of this study can guide future work by supporting researchers select appropriate methods based on their specific characteristics and the scope of the study, particularly in efforts to better understand human behaviors relevant to transmission of infectious diseases. A key consideration for future work is the practicality of large-scale sensor deployment, particularly in ensuring participant compliance and effective monitoring across geographically dispersed households.

## List of abbreviations

**Abbreviations Meaning**

LMICs: Low- and middle-income countries
ID: Identification number
RSSI: Received signal strength indicator
SD: Standard deviation

## Declarations

### Consent for publication

Not applicable

### Availability of data and materials

The datasets generated and/or analyzed during the current study are available in the GlobalMix-sensor-analysis repository, https://github.com/lopmanlab/GlobalMix-sensor-analysis.git

### Author’s contributions

M.S. led the data analysis and manuscript writing. M.S. and M. Kiti were responsible for data cleaning and management. M. Kiti also contributed substantially to manuscript revisions. Authors 2-22 (M. Kiti through K.N.) contributed to the design and implementation of the study. Authors 5, 14-18 (C.S., H.M. through R.A.) oversaw participant recruitment and data collection at their respective local sites. Authors 5-18 (C.S. through R.A.) contributed to data collection and field coordination at their respective local sites. B.L. and S.O., as principal investigators of the overarching GlobalMix study, provided strategic oversight and critical feedback on the manuscript. K.N. supervised the development of this manuscript, providing regular guidance and critical feedback throughout. All authors reviewed the manuscript, provided important intellectual input, and approved the final version.

## Acknowledgements

Not applicable

