## Supplemental materials for "Comparison of contact diaries and wearable proximity sensors in measuring household contacts in low- and middle-income countries"

Supplemental Table 1: Results of logistic regression analysis

This table is too wide, so it is uploaded in excel format.

Supplemental Figure 1: Flow chart of number of participants in cleaning steps

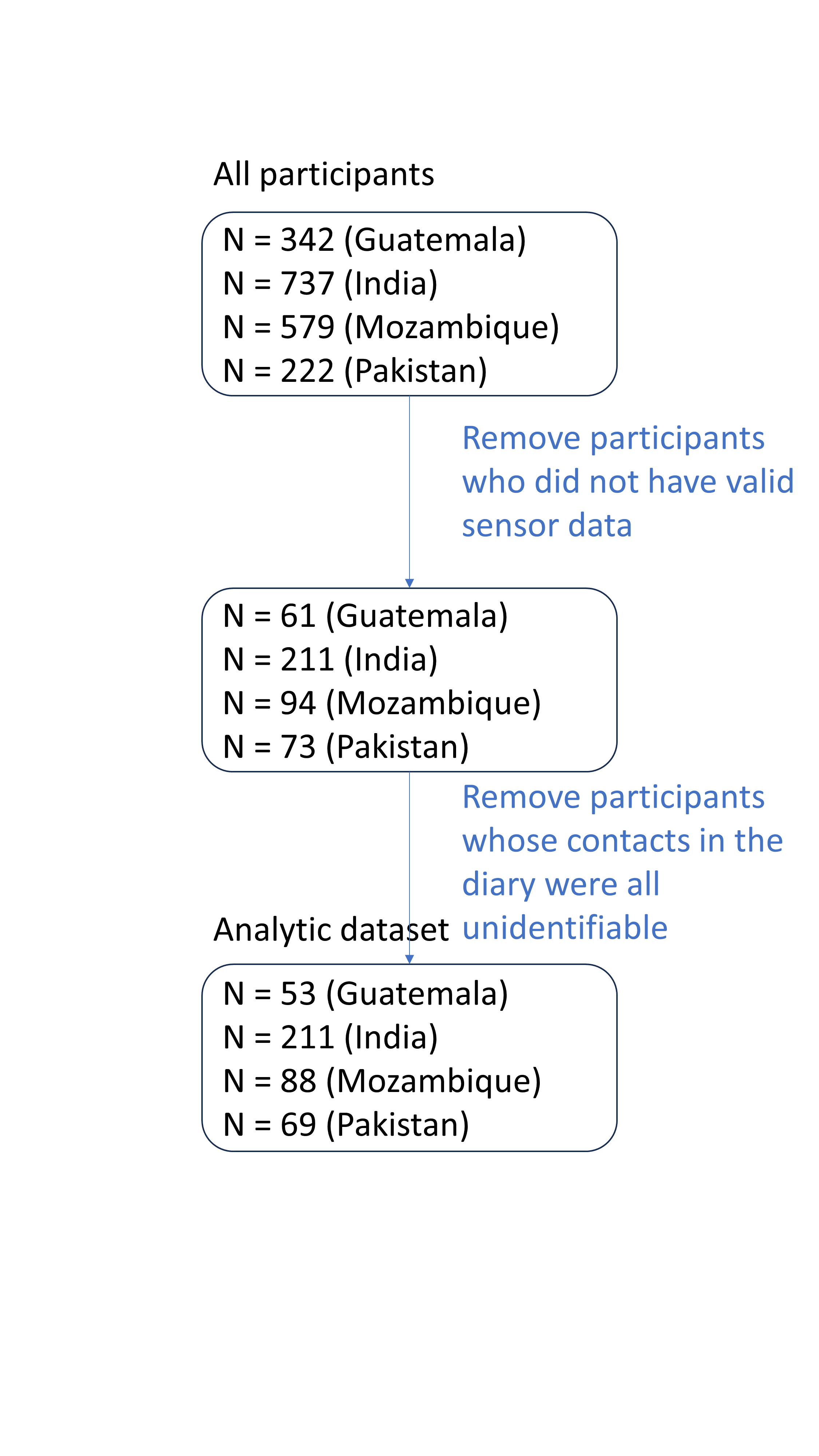

Supplemental Figure 2: Comparison of contacts measured in two methods by contacts’ relationship to participants

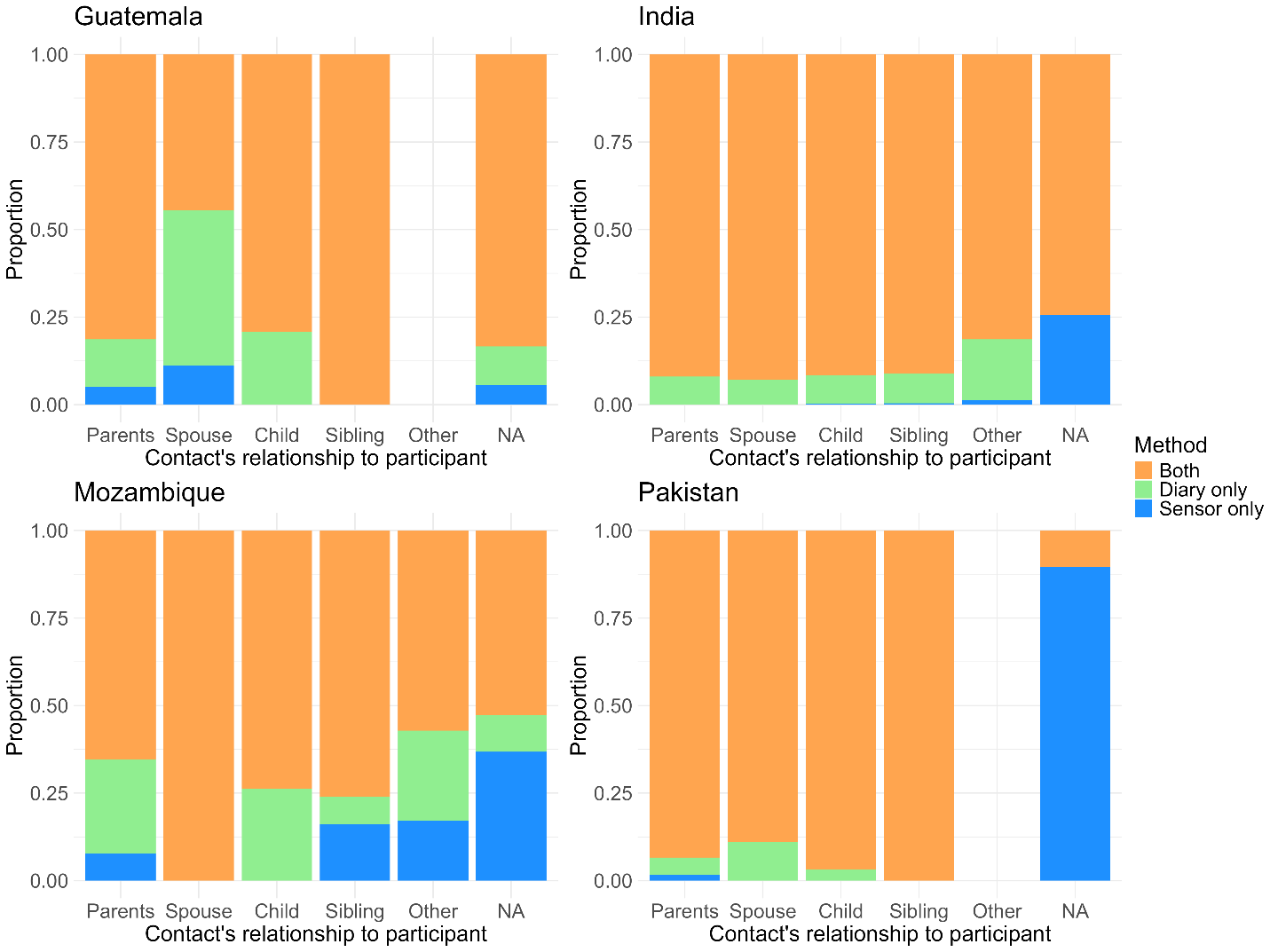

Legend: The figure shows the proportion of the number of contact pairs reported by both methods, diary only, and sensor only by participant age and country. NA means the contacts which household member relationship data is not available.
